# Association of Hemo-Metabolic Trajectory and Cardiogenic Shock Mortality: Analysis from the CSWG Registry

**DOI:** 10.1101/2024.01.05.23300478

**Authors:** Wissam Khalife, Manreet K Kanwar, Jacob Abraham, Kevin John, Aiham Albaeni, Borui Li, Yijing Zhang, Van-Khue Ton, Maya Guglin, Arthur R Garan, Rachna Kataria, Vanessa Blumer, Gavin W Hickey, Song Li, Saraschandra Vallabhajosyula, Shashank S Sinha, Jaime Hernandez-Montfort, Elric Zweck, Chloe Kong, MaryJane Farr, Justin Fried, Shelley Hall, Neil M Harwani, Claudius Mahr, Sandeep Nathan, Paavni Sangal, Andrew Schwartzman, Arvind Bhimaraj, Ju Kim, Alec A Vishnevsky, Esther Vorovich, Karol D. Walec, Peter Zazzali, Mohit Pahuja, Daniel Burkhoff, Navin K Kapur

## Abstract

**Background:** Cardiogenic shock (CS) is as a hemodynamic disorder that can progress to systemic metabolic derangements. Prior studies have reported hemodynamic parameters associated with mortality in limited cohorts or at single time points. Hemodynamic trajectories have not been described.

**Objectives:** We studied the association between hemodynamics and in-hospital mortality in patients with CS due to heart failure (HF-CS) and acute myocardial infarction (AMI-CS).

**Methods:** Using data from the large multicenter Cardiogenic Shock Working Group (CSWG) registry, we analyzed hemo-metabolic data obtained at the time of pulmonary artery catheter (PAC) insertion (baseline) and at PAC removal or death (final). Univariable regression analyses for prediction of in-hospital mortality were conducted for baseline and final hemo-metabolic values, as well as the interval change (delta-P), and analyzed based on CS etiology and survival status.

**Results:** 2,260 patients with PAC data were included (70% male, age 61±14, 61% HF-CS, 27% AMI-CS). In-hospital mortality was higher in the AMI-CS group (40.1%) compared to HF-CS (22.4%), p<0.001). In the **HF-CS** cohort, survivors exhibited lower right atrial pressure (RAP), pulmonary artery pressures (PAP), cardiac output/index (CO/CI), lactate and higher blood pressure (BP) than non-survivors at baseline. In this cohort, during hospitalization, improvement in metabolic (AST, lactate), BP, hemodynamic (RAP, PAPi, PA compliance for right sided profile and CO/CI for left sided profile), had association with survival. In the **AMI-CS** cohort, a lower systolic BP and higher PAP were associated with odds of death at baseline. Improvement in metabolic (lactate), BP, hemodynamic (RAP, PAPi for right-sided profile and CO/CI for left-sided profile) were associated with survival.

**Conclusions:** In a large contemporary CS registry, few hemo-metabolics at baseline determined survival in AMI-CS; rather hemodynamic trajectories had a strong association with outcomes in both cohorts. These findings suggest the importance of monitoring hemo-metabolic trajectories to tailor management in patients with CS.

**What is New?:**

- While previous cardiogenic shock studies have reported hemodynamic parameters associated with mortality in limited cohorts or at single time points, we used data from a large multi-center registry to analyze hemodynamic trajectory in patients with cardiogenic shock from pulmonary artery catheter insertion to removal.
- We found that few baseline hemodynamic parameters were predictive of survival in AMI-CS. However, in both AMI-CS and HF-CS, the hemodynamic trajectory was strongly associated with outcomes.

**What Are the Clinical Implications?:**

- Our findings suggest that targeted interventions in patients with cardiogenic shock impact clinical outcomes independently of baseline hemodynamic derangement and highlight the importance of invasive hemodynamic monitoring to tailor management in these patients.

## INTRODUCTION

Cardiogenic shock (CS) is a syndrome of primary cardiac dysfunction that results in inadequate cardiac output and hypotension.(1) If inadequately treated, these initial hemodynamic abnormalities result in a complex cascade of tissue hypo-perfusion and systemic injury that precipitate end-organ failure and metabolic acidosis.(2) Clinical studies have demonstrated that therapies targeting hemodynamic disturbances, including acute mechanical circulatory support (MCS) devices, are ineffective if employed non-selectively during late stages of shock, when tissue injury and acidosis are irreversible. The Society for Cardiovascular Angiography and Intervention (SCAI) recently revised a staging scheme of CS severity to reflect the dynamic continuum of hemodynamic and metabolic disturbances that occur during CS and its treatment.(3,4)

Hemodynamic data obtained using a pulmonary artery catheter (PAC) allow for invasive measurement of cardiac filling pressures, measurement of cardiac output (CO) and other parameters.(5) These measures, interpreted together, provide a robust assessment of left and right heart performance that can be used to make prognostic assessments.(6,7) More importantly, PAC is maintained for several days to assess improvement or deterioration along the hemo-metabolic cascade which then impact decisions to escalate or de-escalate MCS therapies. Hemodynamic parameters including RAP, mean arterial pressure (MAP) left ventricular end diastolic pressure (LVEDP), PAPi obtained at baseline have been associated with outcomes in CS.(8–10)

While previous studies have examined the prognostic impact of invasive hemodynamics assessed at a single time point, hemodynamic trajectories in CS during the course of hospitalization and their association with clinical outcomes in real world data have not been described. The insights gained from such an analysis could inform clinicians about which changes in hemodynamic parameters are most strongly associated with outcomes in CS. The aim of this study is to characterize the prognostic significance of hemodynamic parameters and their trajectory using PAC-derived hemodynamics recorded in the Cardiogenic Shock Working Group (CWSG) registry.

## METHODS

### Data Source

The CSWG is an academic research consortium comprised of 34 community and university-affiliated hospitals across the United States. For this analysis, 20 sites contributing registry data between 2016 and 2022 were included. The registry includes a standardized set of data elements (patients, procedural and outcomes) pre-defined by principal investigators and collected retrospectively. CS diagnosis was physician-adjudicated at each site and defined as a sustained episode of one out of the following: systolic blood pressure (SBP) < 90 mmHg for at least 30 minutes; use of vasoactive agents; a CI <2.2 L/min/m^2^ in the absence of hypovolemia determined to be secondary to cardiac dysfunction; or use of one or more temporary MCS devices for clinically-suspected CS. Treatments for CS were left to the discretion of the clinicians at each center and were not guided by a prescribed algorithm.

Patient laboratory and hemodynamic data were collected at multiple time points, including closest to time of admission (baseline), at PAC insertion, at PAC removal or death. The PAC-derived hemodynamic parameters include both right heart parameters (RAP, pulmonary artery systolic, diastolic and mean pressure or PASP, PADP and mPAP respectively, SVO2) and left heart parameters (PCWP, cardiac output and cardiac index; CO and CI, respectively). Secondary hemodynamic parameters (e.g. transpulmonary gradient or TPG, diastolic pulmonary gradient or DPG, PAPi, aortic power index (API), RAP/ PCWP ratio, CPO, right ventricular stroke work index RVSWI etc.) were calculated from the collected data, as possible (Supplement Table 1). Additional data, including vital signs (SBP, mean arterial pressure or MAP and heart rate or HR) as well as laboratory data (including lactate, pH, metabolic profile etc.) were also collected. The SCAI shock stage was retrospectively assigned as previously described.(11) Patient outcomes were reported as survival at the time of hospital discharge including native heart survival or heart replacement therapies (durable LVAD or heart transplantation). Quality assurance was achieved through adjudication at each site by the respective clinical coordinators and principal investigator. Values were centrally audited and screened by the CSWG research team for any discrepancies or major outliers and resolved with submitting site.

### Study Population

Between 2016 and 2022, data from 2,260 CS patients with PAC were collected. CS etiology was reported by each site as either due to AMI or HF. AMI-CS was defined as any primary diagnosis of either non-ST-segment elevation (NSTEMI) or ST-segment elevation (STEMI). HF-CS was defined as any primary diagnosis of acute on chronic HF, or de novo HF, not otherwise related to AMI. Patients with post-cardiotomy CS, primary RV shock or unspecified etiology were considered as “others”.

### Statistical Analysis

Baseline clinical characteristics for HF-CS and AMI-CS cohorts were reported as percentages for categorical variables and as means and standard deviation for continuous variables. The hemodynamic parameters were reported as means and standard deviation at PAC insertion (as **baseline dataset**) and PAC removal or patient death (**final dataset**), stratified by etiology of CS (HF-CS vs AMI-CS) and by survival status at hospital discharge (survivor vs non-survivor). The change in each hemodynamic parameter from PAC insertion to PAC removal, collectively referred to as the ‘**delta P**’ (change in parameter), was also similarly reported. If a patient went on receive LVAD or heart transplantation, final dataset was counted as the last dataset available prior to surgery. For continuous variables with normal distributions, the p-values were calculated using the student’s t-test, and for variables with skewed distributions, the p-values were calculated using the Wilcoxon test. Categorical variables were expressed as frequency and compared using chi-square tests of independence. Univariable logistic regression was performed for each hemodynamic and metabolic variable for mortality association. Odds ratios, 95% CI, and p values were reported. Statistical analyses were preformed using SAS 9.4, p values <0.05 were considered significant for all statistical tests.

## RESULTS

### Baseline characteristics

During the study period, the CSWG registry included data from 3,524 patients with CS of whom, 2,260 (64%) patients underwent PAC placement and constitutes the study population for this analysis. These include 1,386 (61%) with HF-CS; 604 (27%) with AMI-CS patients and 270 patients classified as ‘others’ (Figure 1 Supplement). Patient characteristics of the AMI-CS and HF-CS study populations are summarized in **Table 1**. Compared to the AMI-CS cohort, patients in the HF-CS cohort were younger (mean age 60.1 vs. 66 years, p<0.001) more likely to be black (20.6% vs. 7.1%, p<0.001), have a history of atrial fibrillation/flutter, chronic kidney disease, valvular disease, and prior CABG (all p<0.001).

**Figure 1:**
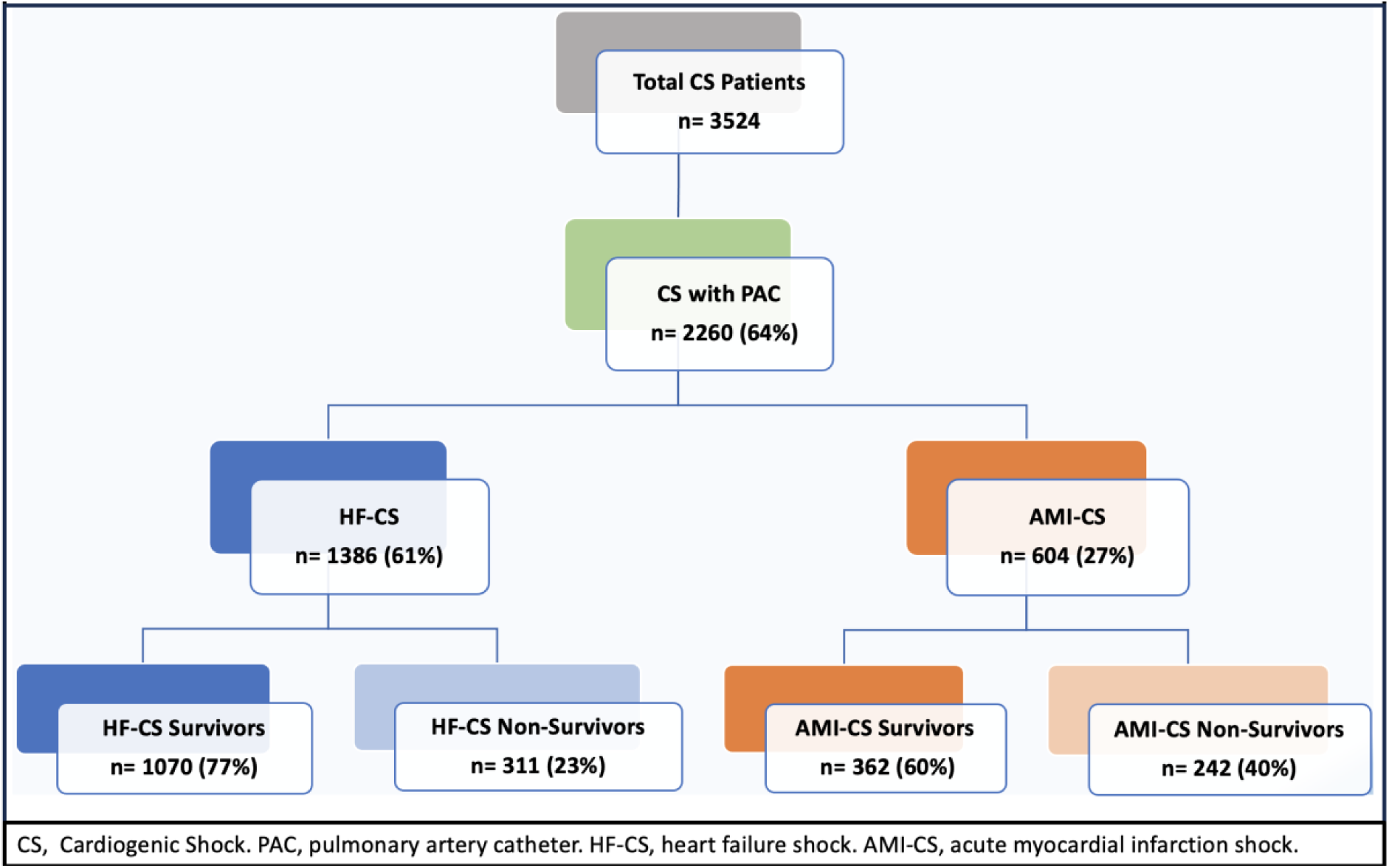
Diagram of the cardiogenic shock patients with pulmonary artery catheter by etiology and survival status.

**Figure 2:**
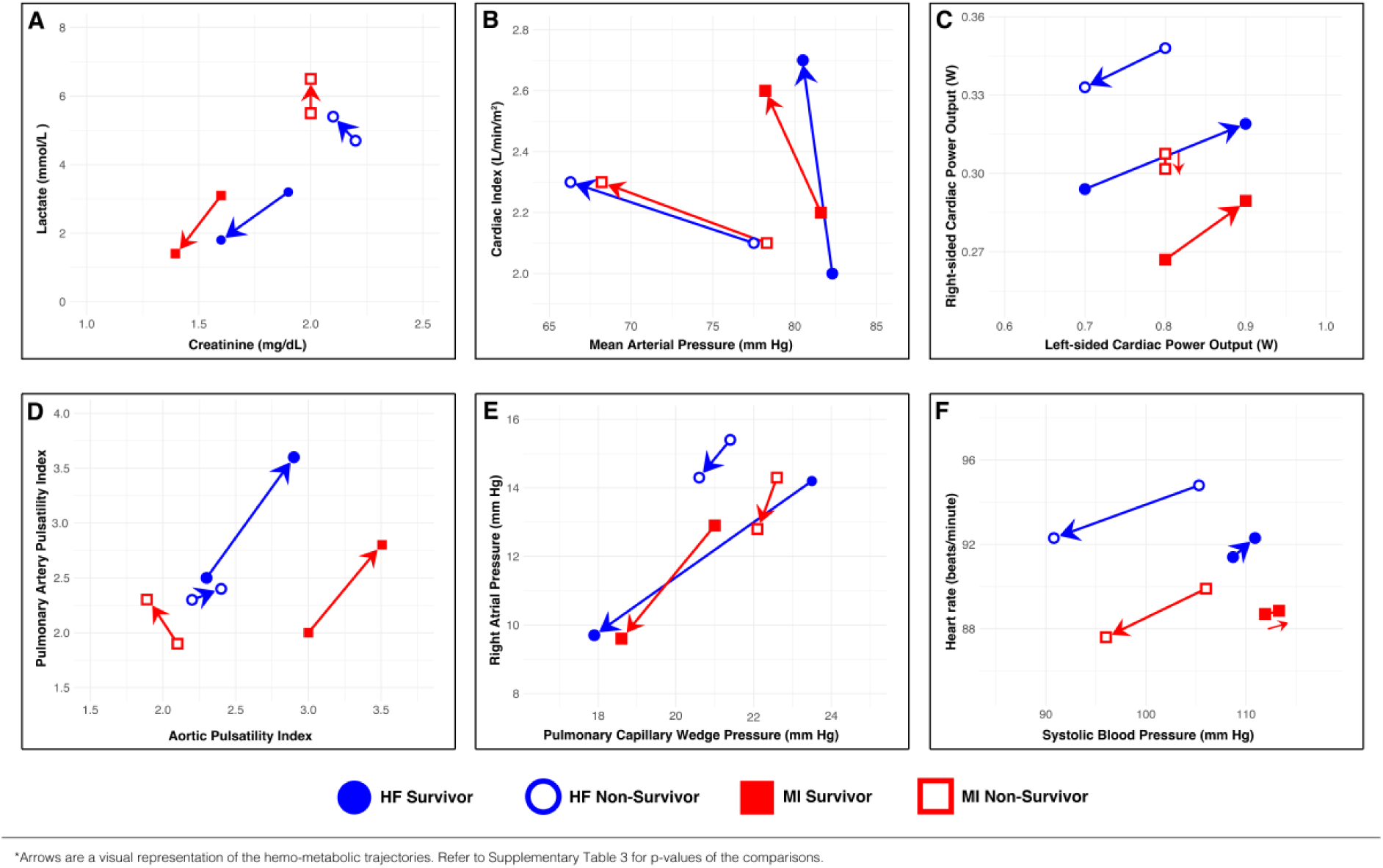
Hemo-metabolic trajectories in AMI-cardiogenic shock and HF-cardiogenic shock. The trajectories of important metabolic and hemodynamic parameters in cardiogenic shock related to acute myocardial infarction (AMI-CS) and cardiogenic shock related to heart failure (HF-CS). Blue full circles represent CS-HF survivors, blue empty circles for HF-CS non-survivors, red full squares for AMI-CS survivors, red empty squares for AMI-CS non-survivors.

**Table 1:**
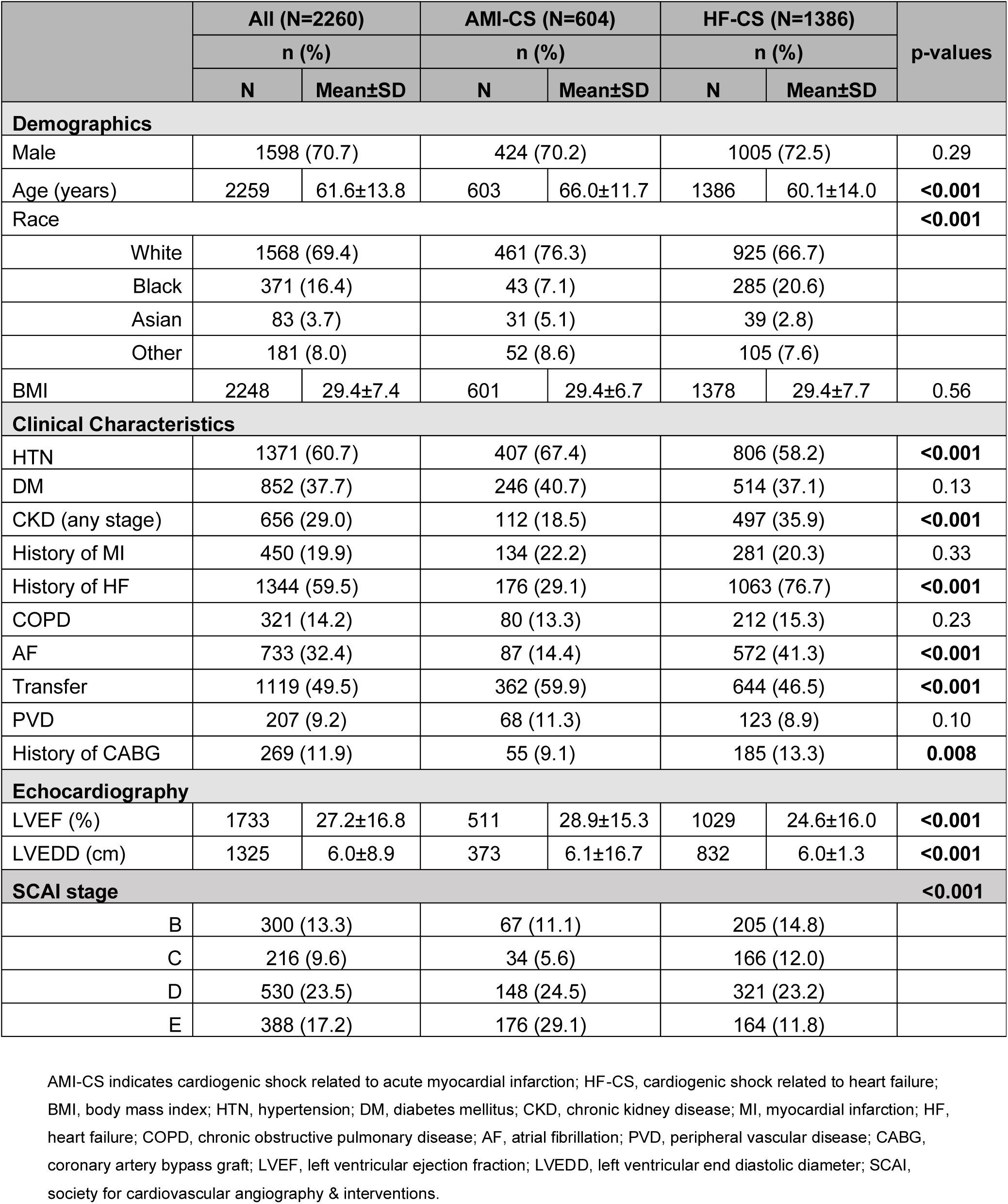
Baseline characteristics of patient population by cardiogenic shock etiology. Patients with shock due to causes other than HF and AMI are not summarized as a separate group due to the small number (n=270).

|  | All (N=2260) |  | AMI-CS (N=604) |  | HF-CS (N=1386) |  | p-values |
| --- | --- | --- | --- | --- | --- | --- | --- |
|  | n (%) |  | n (%) |  | n (%) |  |  |
|  | N | Mean±SD | N | Mean±SD | N | Mean±SD |  |
| Demographics |  |  |  |  |  |  |  |
| Male | 1598 (70.7) |  | 424 (70.2) |  | 1005 (72.5) |  | 0.29 |
| Age (years) | 2259 | 61.6±13.8 | 603 | 66.0±11.7 | 1386 | 60.1±14.0 | <0.001 |
| Race |  |  |  |  |  |  | <0.001 |
| White | 1568 (69.4) |  | 461 (76.3) |  | 925 (66.7) |  |  |
| Black | 371 (16.4) |  | 43 (7.1) |  | 285 (20.6) |  |  |
| Asian | 83 (3.7) |  | 31 (5.1) |  | 39 (2.8) |  |  |
| Other | 181 (8.0) |  | 52 (8.6) |  | 105 (7.6) |  |  |
| BMI | 2248 | 29.4±7.4 | 601 | 29.4±6.7 | 1378 | 29.4±7.7 | 0.56 |
| Clinical Characteristics |  |  |  |  |  |  |  |
| HTN | 1371 (60.7) |  | 407 (67.4) |  | 806 (58.2) |  | <0.001 |
| DM | 852 (37.7) |  | 246 (40.7) |  | 514 (37.1) |  | 0.13 |
| CKD (any stage) | 656 (29.0) |  | 112 (18.5) |  | 497 (35.9) |  | <0.001 |
| History of MI | 450 (19.9) |  | 134 (22.2) |  | 281 (20.3) |  | 0.33 |
| History of HF | 1344 (59.5) |  | 176 (29.1) |  | 1063 (76.7) |  | <0.001 |
| COPD | 321 (14.2) |  | 80 (13.3) |  | 212 (15.3) |  | 0.23 |
| AF | 733 (32.4) |  | 87 (14.4) |  | 572 (41.3) |  | <0.001 |
| Transfer | 1119 (49.5) |  | 362 (59.9) |  | 644 (46.5) |  | <0.001 |
| PVD | 207 (9.2) |  | 68 (11.3) |  | 123 (8.9) |  | 0.10 |
| History of CABG | 269 (11.9) |  | 55 (9.1) |  | 185 (13.3) |  | 0.008 |
| Echocardiography |  |  |  |  |  |  |  |
| LVEF (%) | 1733 | 27.2±16.8 | 511 | 28.9±15.3 | 1029 | 24.6±16.0 | <0.001 |
| LVEDD (cm) | 1325 | 6.0±8.9 | 373 | 6.1±16.7 | 832 | 6.0±1.3 | <0.001 |
| SCAI stage |  |  |  |  |  |  | <0.001 |
| B | 300 (13.3) |  | 67 (11.1) |  | 205 (14.8) |  |  |
| C | 216 (9.6) |  | 34 (5.6) |  | 166 (12.0) |  |  |
| D | 530 (23.5) |  | 148 (24.5) |  | 321 (23.2) |  |  |
| E | 388 (17.2) |  | 176 (29.1) |  | 164 (11.8) |  |  |

AMI-CS indicates cardiogenic shock related to acute myocardial infarction; HF-CS, cardiogenic shock related to heart failure; BMI, body mass index; HTN, hypertension; DM, diabetes mellitus; CKD, chronic kidney disease; MI, myocardial infarction; HF, heart failure; COPD, chronic obstructive pulmonary disease; AF, atrial fibrillation; PVD, peripheral vascular disease; CABG, coronary artery bypass graft; LVEF, left ventricular ejection fraction; LVEDD, left ventricular end diastolic diameter; SCAI, society for cardiovascular angiography & interventions.

### Hemo-metabolic characteristics

Compared to the AMI-CS cohort, patients in the HF-CS cohort were more likely to present with a higher heart rate (95 vs 89 bpm, p=0.01), serum creatinine (2.0 vs 1.7 mg/dL, p<0.001) and bicarbonate (23.5 vs 21.7 mg/dL, p<0.001), but a lower lactate (3.9 vs. 4.7 mmol/L p<0.03). **(Table 2)** For the left-sided hemodynamics standpoint, patients in HF-CS had a higher PCWP (23.2 vs. 21.5 mmHg, p<0.001), PADP (26.4 vs. 23.1 mmHg, p<0.001) and lower CI (2.0 vs 2.2 L/min/m^2^, p=0.01) at baseline, compared to AMI-CS. (Supplement Table 2) For the right-sided hemodynamic parameters, HF-CS pts had a higher RAP, PAP and PAPi compared to MI-CS at baseline (all p<0.01). (Table 2) When comparing the baseline and final dataset for the AMI-CS cohort, CO, CI, PAPi and RVSWi increased while RAP and PAP decreased significantly. For the HF-CS cohort, the final dataset had a significantly lower PCWP, PAP, RAP and a higher CO/CI, CPO, PAPi and RVSWi compared to baseline.

**Table 2:**
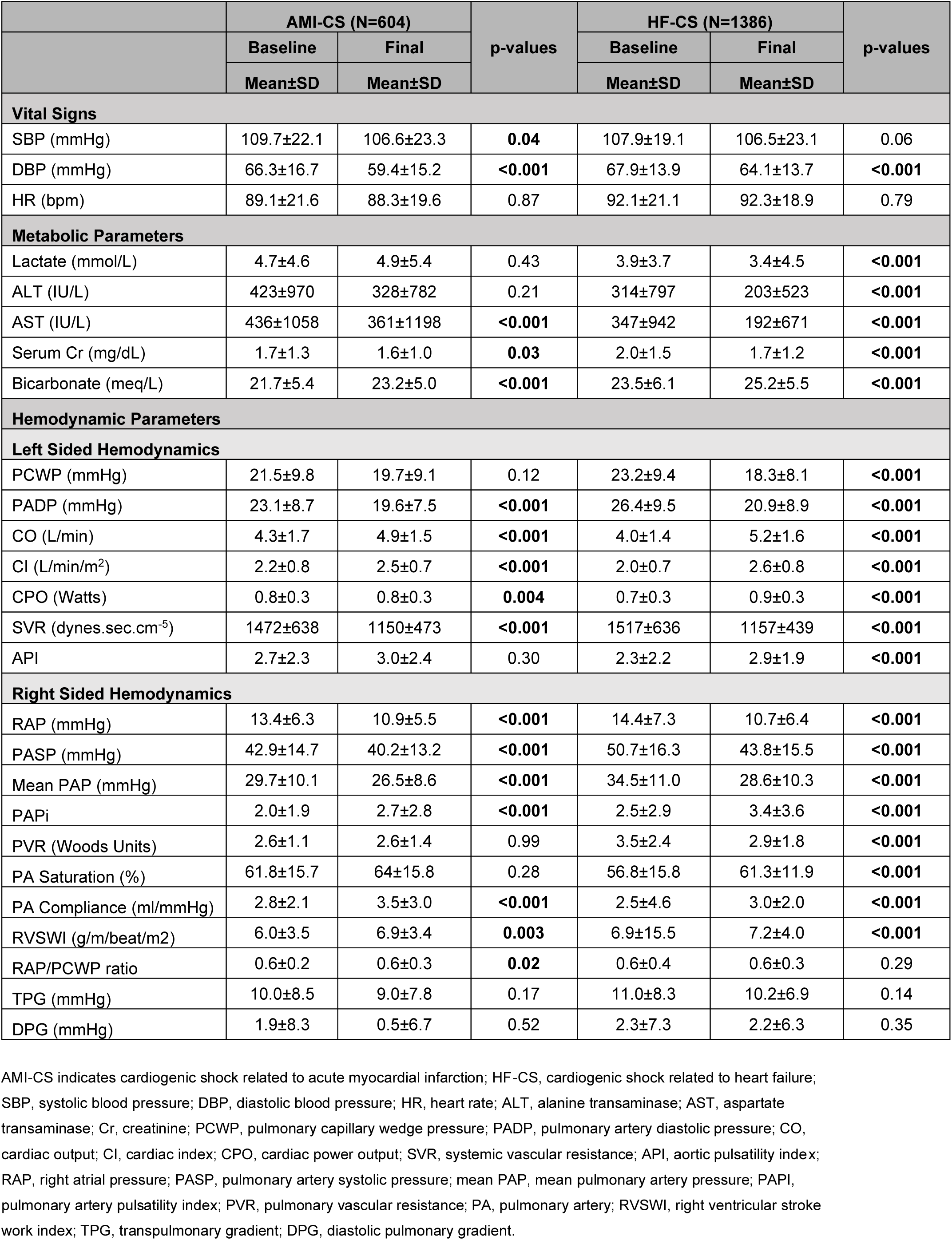
Baseline and final hemo-metabolic variables in the AMI-CS and HF-CS.

|  | AMI-CS (N=604) |  | p-values | HF-CS (N=1386) |  | p-values |
| --- | --- | --- | --- | --- | --- | --- |
|  | Baseline | Final |  | Baseline | Final |  |
|  | Mean±SD | Mean±SD |  | Mean±SD | Mean±SD |  |
| Vital Signs |  |  |  |  |  |  |
| SBP (mmHg) | 109.7±22.1 | 106.6±23.3 | 0.04 | 107.9±19.1 | 106.5±23.1 | 0.06 |
| DBP (mmHg) | 66.3±16.7 | 59.4±15.2 | <0.001 | 67.9±13.9 | 64.1±13.7 | <0.001 |
| HR (bpm) | 89.1±21.6 | 88.3±19.6 | 0.87 | 92.1±21.1 | 92.3±18.9 | 0.79 |
| Metabolic Parameters |  |  |  |  |  |  |
| Lactate (mmol/L) | 4.7±4.6 | 4.9±5.4 | 0.43 | 3.9±3.7 | 3.4±4.5 | <0.001 |
| ALT (IU/L) | 423±970 | 328±782 | 0.21 | 314±797 | 203±523 | <0.001 |
| AST (IU/L) | 436±1058 | 361±1198 | <0.001 | 347±942 | 192±671 | <0.001 |
| Serum Cr (mg/dL) | 1.7±1.3 | 1.6±1.0 | 0.03 | 2.0±1.5 | 1.7±1.2 | <0.001 |
| Bicarbonate (meq/L) | 21.7±5.4 | 23.2±5.0 | <0.001 | 23.5±6.1 | 25.2±5.5 | <0.001 |
| Hemodynamic Parameters |  |  |  |  |  |  |
| Left Sided Hemodynamics |  |  |  |  |  |  |
| PCWP (mmHg) | 21.5±9.8 | 19.7±9.1 | 0.12 | 23.2±9.4 | 18.3±8.1 | <0.001 |
| PADP (mmHg) | 23.1±8.7 | 19.6±7.5 | <0.001 | 26.4±9.5 | 20.9±8.9 | <0.001 |
| CO (L/min) | 4.3±1.7 | 4.9±1.5 | <0.001 | 4.0±1.4 | 5.2±1.6 | <0.001 |
| CI (L/min/m²) | 2.2±0.8 | 2.5±0.7 | <0.001 | 2.0±0.7 | 2.6±0.8 | <0.001 |
| CPO (Watts) | 0.8±0.3 | 0.8±0.3 | 0.004 | 0.7±0.3 | 0.9±0.3 | <0.001 |
| SVR (dynes.sec.cm⁻⁵) | 1472±638 | 1150±473 | <0.001 | 1517±636 | 1157±439 | <0.001 |
| API | 2.7±2.3 | 3.0±2.4 | 0.30 | 2.3±2.2 | 2.9±1.9 | <0.001 |
| Right Sided Hemodynamics |  |  |  |  |  |  |
| RAP (mmHg) | 13.4±6.3 | 10.9±5.5 | <0.001 | 14.4±7.3 | 10.7±6.4 | <0.001 |
| PASP (mmHg) | 42.9±14.7 | 40.2±13.2 | <0.001 | 50.7±16.3 | 43.8±15.5 | <0.001 |
| Mean PAP (mmHg) | 29.7±10.1 | 26.5±8.6 | <0.001 | 34.5±11.0 | 28.6±10.3 | <0.001 |
| PAPi | 2.0±1.9 | 2.7±2.8 | <0.001 | 2.5±2.9 | 3.4±3.6 | <0.001 |
| PVR (Woods Units) | 2.6±1.1 | 2.6±1.4 | 0.99 | 3.5±2.4 | 2.9±1.8 | <0.001 |
| PA Saturation (%) | 61.8±15.7 | 64±15.8 | 0.28 | 56.8±15.8 | 61.3±11.9 | <0.001 |
| PA Compliance (ml/mmHg) | 2.8±2.1 | 3.5±3.0 | <0.001 | 2.5±4.6 | 3.0±2.0 | <0.001 |
| RVSWI (g/m/beat/m2) | 6.0±3.5 | 6.9±3.4 | 0.003 | 6.9±15.5 | 7.2±4.0 | <0.001 |
| RAP/PCWP ratio | 0.6±0.2 | 0.6±0.3 | 0.02 | 0.6±0.4 | 0.6±0.3 | 0.29 |
| TPG (mmHg) | 10.0±8.5 | 9.0±7.8 | 0.17 | 11.0±8.3 | 10.2±6.9 | 0.14 |
| DPG (mmHg) | 1.9±8.3 | 0.5±6.7 | 0.52 | 2.3±7.3 | 2.2±6.3 | 0.35 |

AMI-CS indicates cardiogenic shock related to acute myocardial infarction; HF-CS, cardiogenic shock related to heart failure; SBP, systolic blood pressure; DBP, diastolic blood pressure; HR, heart rate; ALT, alanine transaminase; AST, aspartate transaminase; Cr, creatinine; PCWP, pulmonary capillary wedge pressure; PADP, pulmonary artery diastolic pressure; CO, cardiac output; CI, cardiac index; CPO, cardiac power output; SVR, systemic vascular resistance; API, aortic pulsatility index; RAP, right atrial pressure; PASP, pulmonary artery systolic pressure; mean PAP, mean pulmonary artery pressure; PAPI, pulmonary artery pulsatility index; PVR, pulmonary vascular resistance; PA, pulmonary artery; RVSWI, right ventricular stroke work index; TPG, transpulmonary gradient; DPG, diastolic pulmonary gradient.

### PAC use and in-hospital clinical outcomes

Median time to PAC insertion was 0.9 days; it was inserted later (1.1 vs 0.4 days, p<0.0001) but maintained for a longer duration in patients with HF-CS than AMI-CS (6.3 vs 4.7 days, p<0.0001). Median time to the final hemodynamic dataset was

8.15 (IQR 4.02-15.97) days for the total cohort and 7.42 (IQR 3.04-13.97) days in those who died. The use of vasoactive drugs throughout hospitalization was similar between HF-CS and AMI-CS. HF-CS patients were less likely than AMI-CS patients to be treated with temporary MCS (IABP, Impella CP, or multiple devices) (53.9% vs 78.8%, p<0.001). Overall, in-hospital mortality in the overall cohort was 27.8% (n=553); 22.4% in HF-CS patients (n=311) and 40.1% in AMI-CS patients (n=242) **(Table 3).** Heart replacement therapy occurred in 371 (26.8%) of patients in the HF-CS group compared to 37 (6.1%) in AMI-CS group (p <0.0001).

**Table 3:** In hospital outcomes and interventions in AMI-CS and HF-CS patients.

|  | All<br>(N=2260) | AMI-CS<br>(n=604) | HF-CS<br>(n=1386) | p-values |
| --- | --- | --- | --- | --- |
|  | N (%) |  |  |  |
| PAC utilization; Median (IQR) |  |  |  |  |
| Admission to PAC insertion (days) | 0.9 (2.9) | 0.4 (1.6) | 1.1 (3.6) | <0.0001 |
| Duration of PAC (days) | 5.8 (9.6) | 4.7 (7.3) | 6.3 (12.2) | <0.0001 |
| In-hospital Interventions |  |  |  |  |
| PCI done at stay | 359 (15.5) | 271 (50.3) | 61 (4.5) | <0.0001 |
| CABG done at stay | 178 (7.9) | 90 (15.3) | 38 (2.8) | <0.0001 |
| MCS devices |  |  |  |  |
| MCS device/s during hospitalization | 1401 (61.9) | 476 (78.8) | 748 (53.9) | <0.0001 |
| IABP alone | 556 (24.6) | 170 (28.1) | 329 (23.7) | 0.04 |
| Impella CP alone | 151 (6.6) | 83 (13.7) | 59 (4.3) | <0.0001 |
| Impella 5/5.5 alone | 100 (4.4) | 15 (2.5) | 81 (5.8) | 0.001 |
| ECMO alone | 111 (4.9) | 21 (3.5) | 48 (3.5) | 0.99 |
| Other/Multiple | 483 (21.3) | 187 (30.9) | 231 (16.7) | <0.0001 |
| MCS devices before PAC insertion | 745 (33) | 197 (32.6) | 437 (31.5) | 0.31 |
| Vasopressors/Inotropes |  |  |  |  |
| Vasopressors/Inotropes during hospitalization | 1903 (84.3) | 526 (87.1) | 1153 (83.2) | 0.70 |
| Vasopressors/inotropes at PAC insertion |  |  |  | 0.001 |
| 0 | 727 (32.2) | 158 (26.2) | 479 (34.6) |  |
| 1 | 687 (30.8) | 143 (23.7) | 481 (34.7) |  |
| 2+ | 433 (19.2) | 124 (20.5) | 251 (18.1) |  |
| Vasopressors/inotropes at PAC removal |  |  |  | <0.0001 |
| 0 | 869 (38.5) | 232 (38.4) | 537 (38.7) |  |
| 1 | 780 (34.5) | 168 (27.8) | 529 (38.2) |  |
| 2+ | 587 (26) | 199 (33) | 305 (22) |  |
| In-Hospital Outcomes |  |  |  |  |
| Cardiac Arrest | 472 (20.8) | 212 (35.1) | 183 (13.2) | <0.0001 |
| Heart Replacement Therapy (LVAD/HT) | 438 (19.6) | 37 (6.1) | 371 (26.8) | <0.0001 |
| Native Heart Recovery | 1168 (51.7) | 324 (53.6) | 698 (50.4) | 0.2 |
| In-hospital mortality | 649 (28.7) | 242 (40.1) | 311 (22.4) | <0.0001 |

AMI-CS indicates cardiogenic shock related to acute myocardial infarction; HF-CS, cardiogenic shock related to heart failure; PAC, pulmonary artery catheter; PCI, percutaneous coronary intervention; CABG, coronary artery bypass graft; MCS, mechanical circulatory support; IABP, intra-aortic balloon pump; ECMO, extracorporeal membrane oxygenation; LAVD, left ventricular assist device; HT, heart transplant.

### Hemodynamic profiles and survival

In unadjusted analysis (**Table 4**), survivors in the HF-CS cohort exhibited lower HR, RAP, PAP, CO, CI, TPG and higher BP (systolic, diastolic, and mean) than non-survivors at baseline. At the time of final dataset, survivors had a lower PAP, PCWP and RAP and a higher BP, CO, CI, CPO, PAPi and PA sat compared to non-survivors. Metabolic markers of lactate, BUN and creatinine were higher for non-survivors, both at baseline and final dataset. During the course of hospitalization, significant improvements from baseline to PAC removal were observed in BP, filling pressures, and CO amongst survivors. (**Supplement Table 3 and 4, Figure 1**) For example, for a 1 mm Hg drop in RAP between baseline and final dataset, there was 0.06 lower odds of mortality; while for every 1 L/min/m^2^ increase in CI, the odds of survival improved by 0.44. (**Supplement Table 5**)

**Table 4:**
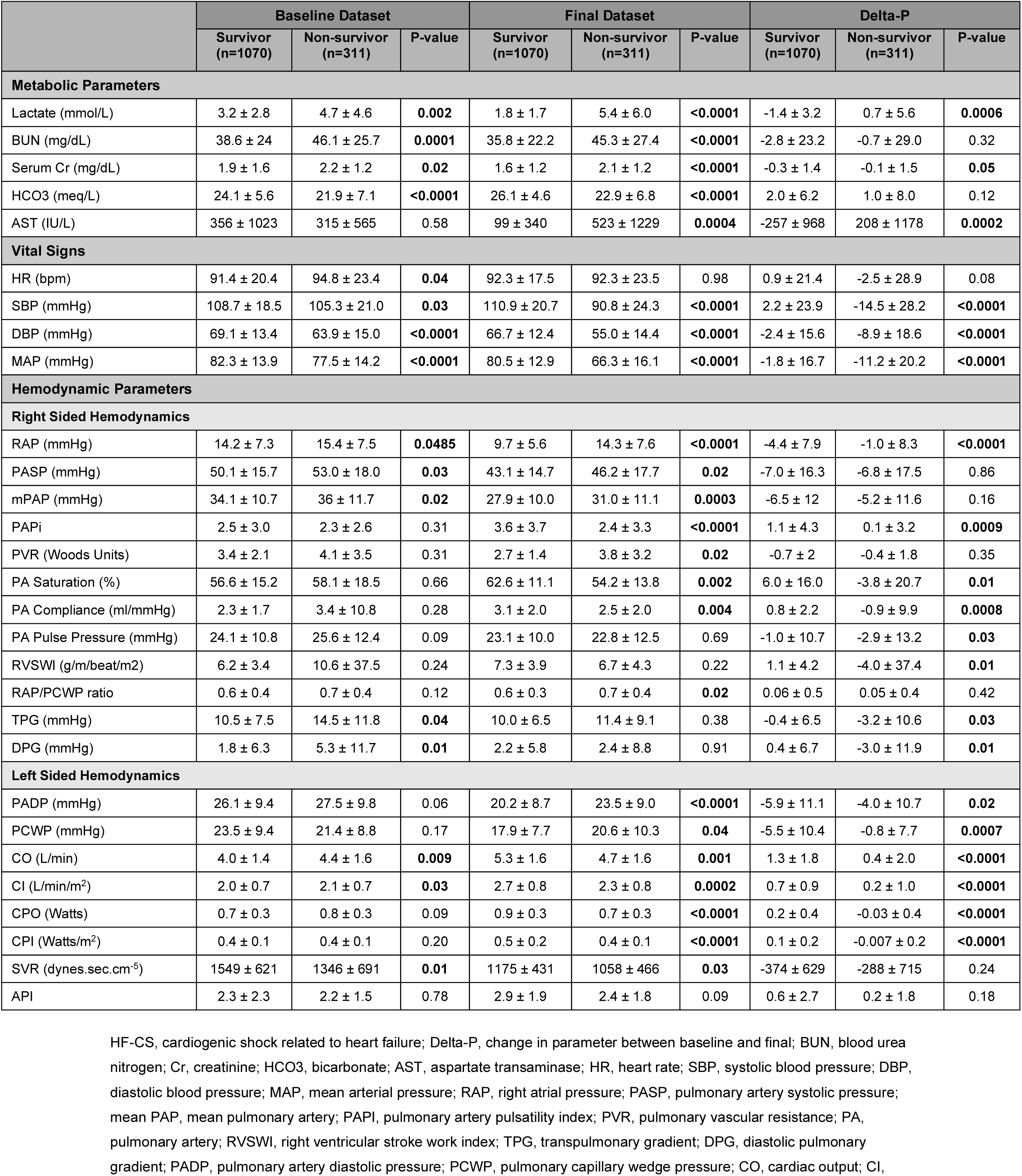
Univariable analysis of hemo-metabolic profile in the HF-CS cohort comparing survivors and non-survivors, at baseline and final dataset.

In the AMI-CS cohort a lower SBP and higher PAP were associated with odds of death at baseline. (**Table 5**) For the final dataset, markers that had the greatest influence on mortality included lower BP, CO, CI and a higher RAP and PAP. During the course of their hospitalization, significant improvements from baseline to PAC removal were observed in BP and CO/ CI amongst survivors. Metabolic markers of lactate, BUN and creatinine were higher for non-survivors, both at baseline and final dataset

**Table 5:** Univariable analysis of hemo-metabolic profile in the AMI-CS cohort comparing survivors and non-survivors, at baseline and final dataset.

|  | Baseline Dataset |  |  | Final Dataset |  |  | Delta-P |  |  |
| --- | --- | --- | --- | --- | --- | --- | --- | --- | --- |
|  | Survivor<br>(n=362) | Non-survivor<br>(n=242) | P-value | Survivor<br>(n=362) | Non-survivor<br>(n=242) | P-value | Survivor<br>(n=362) | Non-survivor<br>(n=242) | P-value |
| **Metabolic Parameters** |  |  |  |  |  |  |  |  |  |
| Lactate (mmol/L) | 3.1 ± 2.6 | 5.5 ± 5.1 | **0.016** | 1.4 ± 0.3 | 6.5 ± 5.8 | **<0.0001** | -1.7 ± 2.6 | 1.1 ± 5.9 | **0.0009** |
| BUN (mg/dL) | 28.0 ± 18.9 | 36.4 ± 20.8 | **0.0001** | 30.6 ± 19.1 | 43.2 ± 25.6 | **<0.0001** | 2.6 ± 19.0 | 6.7 ± 28.3 | 0.13 |
| Serum Cr (mg/dL) | 1.6 ± 1.1 | 2.0 ± 1.5 | **0.002** | 1.4 ± 0.9 | 2.0 ± 1.1 | **<0.0001** | -0.2 ± 0.9 | -0.01 ± 1.6 | 0.23 |
| HCO<sub>3</sub> (meq/L) | 22.1 ± 5.3 | 21.1 ± 5.6 | 0.12 | 24.6 ± 3.9 | 21.4 ± 5.7 | **<0.0001** | 2.4 ± 6.2 | 0.3 ± 7.0 | **0.007** |
| AST (IU/L) | 380 ± 827 | 517 ± 1331 | 0.47 | 107 ± 211 | 734 ± 1812 | **0.008** | -273 ± 848 | 217 ± 2181 | 0.09 |
| **Vital Signs** |  |  |  |  |  |  |  |  |  |
| HR (bpm) | 88.7 ± 21.5 | 89.8 ± 21.7 | 0.62 | 88.8 ± 16.9 | 87.6 ± 23.4 | 0.60 | 0.1 ± 23.8 | -2.2 ± 28.5 | 0.41 |
| SBP (mmHg) | 111.9 ± 22.1 | 106 ± 21.7 | **0.01** | 113 ± 18.9 | 96.0 ± 26.0 | **<0.0001** | 1.2 ± 27.3 | -10.0 ± 31.5 | **0.0003** |
| DBP (mmHg) | 67.0 ± 16.8 | 65.2 ± 16.6 | 0.32 | 61.9 ± 14.0 | 55.4 ± 16.2 | **0.0001** | -5.1 ± 17.7 | -9.8 ± 22.2 | **0.03** |
| MAP (mmHg) | 81.6 ± 15.9 | 78.3 ± 15.1 | 0.05 | 78.2 ± 12.6 | 68.2 ± 16.1 | **<0.0001** | -3.4 ± 18.6 | -10.1 ± 20.9 | **0.002** |
| **Hemodynamic Parameters** |  |  |  |  |  |  |  |  |  |
| **Right Sided Hemodynamics** |  |  |  |  |  |  |  |  |  |
| RAP (mmHg) | 12.9 ± 6.4 | 14.3 ± 5.9 | 0.06 | 9.6 ± 4.7 | 12.8 ± 6.2 | **<0.0001** | -3.3 ± 7.4 | -1.4 ± 7.3 | **0.04** |
| PASP (mmHg) | 41.3 ± 14.0 | 45.4 ± 15.4 | **0.01** | 39.4 ± 12.6 | 41.6 ± 13.9 | 0.13 | -1.9 ± 13.9 | -3.8 ± 15.8 | 0.22 |
| mPAP (mmHg) | 28.4 ± 9.8 | 31.7 ± 10.3 | **0.0006** | 25.4 ± 8 | 28.3 ± 9.1 | **0.004** | -2.6 ± 9.9 | -3.8 ± 11.2 | 0.28 |
| PAPi | 2.0 ± 1.8 | 1.9 ± 2.1 | 0.80 | 3.0 ± 2.6 | 2.3 ± 3.0 | **0.049** | 1.0 ± 3.0 | 0.4 ± 3.6 | 0.13 |
| PVR (Woods Units) | 2.8 ± 1.2 | 2.3 ± 0.9 | 0.32 | 2.8 ± 1.5 | 2.3 ± 0.9 | 0.31 | 0.01 ± 1.3 | -0.04 ± 0.6 | 0.87 |
| PA Saturation (%) | 60.1 ± 14.2 | 64.8 ± 18.0 | 0.32 | 64.8 ± 14.0 | 62.7 ± 19 | 0.67 | 4.7 ± 13.5 | -2.1 ± 17.7 | 0.11 |
| PA Compliance (ml/mmHg) | 2.9 ± 2.0 | 2.5 ± 2.1 | 0.19 | 3.7 ± 3.3 | 3.2 ± 2.3 | 0.22 | 0.8 ± 3.5 | 0.7 ± 2.8 | 0.86 |
| PA Pulse Pressure (mmHg) | 19.6 ± 8.7 | 20.7 ± 10.4 | 0.32 | 21.0 ± 9.1 | 19.9 ± 10.6 | 0.27 | 1.4 ± 10 | -0.8 ± 11 | **0.047** |
| RVSWI (g/m/beat/m<sup>2</sup>) | 6.0 ± 3.7 | 5.8 ± 2.9 | 0.67 | 7.0 ± 3.5 | 6.6 ± 3.0 | 0.38 | 1.0 ± 4.3 | 0.8 ± 3.4 | 0.73 |
| RAP/PCWP ratio | 0.6 ± 0.2 | 0.7 ± 0.3 | 0.65 | 0.6 ± 0.3 | 0.6 ± 0.2 | 0.92 | -0.05 ± 0.3 | -0.1 ± 0.2 | 0.68 |
| TPG (mmHg) | 9.3 ± 6.0 | 11.6 ± 12.1 | 0.45 | 9.9 ± 8.5 | 7.1 ± 6.0 | 0.15 | 0.6 ± 7.2 | -4.5 ± 11.6 | 0.09 |
| DPG (mmHg) | 1.1 ± 5.7 | 3.5 ± 12.2 | 0.43 | 0.4 ± 7.4 | 0.7 ± 5.2 | 0.85 | -0.8 ± 6.5 | -2.8 ± 11.5 | 0.48 |
| **Left Sided Hemodynamics** |  |  |  |  |  |  |  |  |  |
| PADP (mmHg) | 22.1 ± 8.6 | 24.8 ± 8.6 | **0.003** | 18.4 ± 6.9 | 21.7 ± 8.1 | **<0.0001** | -3.7 ± 9.3 | -3.1 ± 10.8 | 0.61 |
| PCWP (mmHg) | 21.0 ± 9.9 | 22.6 ± 10.0 | 0.57 | 18.6 ± 8.3 | 22.1 ± 10.4 | 0.20 | -2.4 ± 9.2 | -0.5 ± 7.5 | 0.41 |
| CO (L/min) | 4.2 ± 1.6 | 4.4 ± 2.0 | 0.63 | 5.1 ± 1.4 | 4.6 ± 1.6 | **0.03** | 0.9 ± 1.9 | 0.2 ± 2.2 | **0.03** |
| CI (L/min/m<sup>2</sup>) | 2.2 ± 0.8 | 2.1 ± 0.9 | 0.84 | 2.6 ± 0.7 | 2.3 ± 0.7 | **0.00** | 0.5 ± 1.0 | 0.1 ± 1.1 | **0.04** |
| CPO (Watts) | 0.8 ± 0.3 | 0.8 ± 0.4 | 0.52 | 0.9 ± 0.3 | 0.8 ± 0.3 | **0.02** | 0.1 ± 0.3 | 0.02 ± 0.4 | 0.25 |
| CPI (Watts/m<sup>2</sup>) | 0.4 ± 0.2 | 0.4 ± 0.2 | 0.12 | 0.5 ± 0.1 | 0.4 ± 0.1 | **0.001** | 0.05 ± 0.2 | 0.02 ± 0.2 | 0.27 |
| SVR (dynes.sec.cm<sup>-5</sup>) | 1496 ± 610 | 1420 ± 693 | 0.49 | 1149 ± 462 | 1152 ± 498 | 0.97 | -347 ± 618 | -268 ± 758 | 0.47 |
| API | 3.0 ± 2.5 | 2.1 ± 1.8 | 0.15 | 3.5 ± 2.6 | 1.9 ± 1.7 | **0.01** | 0.5 ± 2.5 | -0.2 ± 1.3 | 0.21 |

AMI-CS indicates cardiogenic shock related to acute myocardial infarction. The rest of the abbreviations are same as for table

Hemodynamic trajectories and survival:

From a metabolic standpoint, lactate, AST, and creatinine improved over hospital course in both AMI- and HF-CS survivors. (**Figure 1A**) Improving lactate in the delta-P assessment had a significant association with survival amongst the metabolic parameters, regardless of etiology. Hemodynamics reflective of end-organ perfusion (i.e., MAP, CI, CPO), RV function (PAPi) and congestion (CVP, PCWP) demonstrate strikingly different trajectories between survivors and non-survivors in both cohorts. While MAP ranges were similar for survivors at baseline and final dataset, it dropped significantly in non-survivors for both AMI- and HF-CS. (**Figure 1B**) CI increased amongst survivors in both HF-CS and AMI-CS groups along the course of hospitalization. As a result, CPO (product of MAP and CO), increased over the course of hospitalization among survivors while it decreased or was unchanged amongst non-survivors. Notably, HF-CS survivors had a lower baseline CPO compared to non-survivors, whose CPO decreased sharply during the course of hospitalization. (**Figure 1C**) Improvement in PAPi in the delta-P cohort was associated with survival in both AMI- and HF-CS. (**Figure 1D**) Survivors in both CS cohorts had similar final values of PCWP and CVP despite HF-CS patients starting with higher absolute PCWP and CVP. (**Figure 1E**) In the HF-CS cohort, non-survivors had higher RAP at PAC insertion (15.4 vs 14.2 mm Hg, p=0.04) and had less reduction in the same during hospitalization (-1.0 vs -4.4, p<0.0001), compared to survivors. Improvement in BP (systolic, diastolic and MAP) in the delta-P cohort was significantly associated with survival, regardless of etiology. (**Figure 1F)**

## DISCUSSION

In this analysis of real-world data from the CSWG registry, we studied associations among hemodynamic and metabolic parameters obtained at PAC insertion (baseline) and removal (final) and interval change in these parameters (delta-P) with in-hospital mortality. We then compared the hemodynamic profiles and trajectories between survivors and non-survivors using univariable analysis in both AMI- and HF-CS. These analyses support several important observations: (i) Amongst patients with HF-CS, survivors had lower baseline CO/CI, RAP, PAP, lactate and higher BP than non-survivors. During the course of their hospitalization, improvement in metabolic (AST and lactate), BP (systolic, diastolic and MAP), hemodynamic (RAP, PAPi, PA compliance for right-sided profile and CO/CI and CPI for left sided profile) had the highest association with survival (ii) Most invasive hemodynamic parameters were not associated with odds of mortality in AMI-CS at baseline. Improvement in metabolic (lactate), BP (systolic, diastolic and mean), hemodynamic (RAP and PAPi for right-sided profile and CO/CI for left-sided profile) were associated with survival (iii) Hemodynamics indicative of end-organ perfusion and congestion demonstrate strikingly different trajectories between survivors and non-survivors in both cohorts. Our findings suggest that hemodynamically guided interventions in CS impact clinical outcomes independently of baseline hemodynamic risk, hence highlighting the importance of the following hemodynamic trajectories to tailor management. To our knowledge, this is the only analysis of hemodynamic trajectories and their impact on mortality in a cohort of patients with CS due to both HF and AMI.

Early hemodynamic assessment is a cornerstone of the therapeutic approach to CS.(7,11) Recent SCAI staging of CS correlated SCAI stage A (at risk) as cardiac index (CI) ≥2.5 L/min/m^2^ and right atrial pressure (RAP) <10 mm Hg; SCAI stage B (beginning) as CI ≥2.2; and SCAI stage C (Classic) as CI <2.2 L/min/m^2^, pulmonary capillary wedge pressure (PCWP) >15 mm Hg, RAP/PCWP ≥0.8, pulmonary artery Pulsatility index (PAPi) <1.85 and cardiac power output (CPO) ≤0.6 W.(4) In addition, several hemodynamic parameters have been identified previously as strong predictors of in-hospital mortality. The Critical Care Cardiology Trials Network recently reported hemodynamics parameters within 24 hours of admission associated with mortality, which included low MAP, systemic vascular resistance (SVR) and PAPi, along with elevated RAP and RAP/ PCWP ratio.(12) In the SHOCK trial registry of 541 patients with CS, CPO and cardiac power index (CPI) were the strongest independent hemodynamic parameters of in-hospital mortality.(13) A CPO value of ≤ 0.53 Watts predicted in-hospital mortality with a c-index of 0.69 and positive and negative predictive values of 59% and 71%, respectively. A sub study of CardShock that examined those with PAC was also demonstrative of the predictive value of early hemodynamic measurements: CI (OR 0.22, 95% 0.09--0.52), CPI (OR 0.347, 95% 0.20--0.60) and stroke-volume index (OR 0.88, 95% 0.82--0.94) were the strongest predictors of 30-day mortality.(14) In our analysis, baseline left sided CPO was not predictive of survival in either HF- of AMI-CS. However, CPO increased over the course of hospitalization among survivors in HF-CS while it decreased or was unchanged amongst non-survivors.

Mean RAP has been identified as another sensitive hemodynamic marker in CS.(11) A retrospective study of 545 patients with CS found RAP ≥ 14 mmHg to be associated with higher mortality rates and end-organ dysfunction as identified by elevated lactate, creatinine, liver enzymes. An earlier study in patients with acutely decompensated HF who underwent PAC insertion in the ESCAPE (Evaluation Study of Congestive Heart Failure and Pulmonary Artery Catheterization Effectiveness) trial reported that final PCWP and RAP were stronger predictors of 6-month outcomes (death, need for HT or LVAD) than CI. A follow-up ESCAPE sub-analysis identified API as a significant predictor of 6-months outcomes, but not final CI, CPO, or PAPi.(15) We have previously reported that baseline RAP ≥12 mm Hg alone or in combination with PCWP ≥18 mm Hg was associated with higher mortality than isolated elevation in PCWP or normal filling pressures (16,17). While baseline RAP was not predictive of outcome in the current study, final RAP and change in RAP were strong predictors. In fact, survivors in both CS cohorts had similar final values of PCWP and CVP despite HF-CS patients starting with higher absolute PCWP and CVP. Lastly, non-survivors had higher CVP at PAC insertion and had less reduction in PCWP and CVP during hospitalization, compared to survivors. The discrepancy in the present findings from prior studies likely relate to the differences in shock severity and treatment intensity (SCAI A in ESCAPE versus SCAI C-E in CSWG) and the larger numbers of patients enrolled in the most recent version of the CSWG registry employed in the current study.

Another important hemodynamic indicator of RV failure in patients with acute inferior MI is PAPi.(18,19) A PAPi value of ≤ 0.9 carries a specificity of 98.3% and a sensitivity of 100% to predict in-hospital mortality in patients with inferior MI undergoing emergent PCI. By contrast, a PAPi value of < 1.85 predicts RV failure after isolated LVAD implantation with 94% sensitivity and 81% specificity.(20) In our analysis, baseline PAPi was not predictive of survival in either etiology by an improvement in PAPi was significantly associated with survival in the HF-CS. This may reflective of the higher rates of MCS utilization and longer duration of PAC monitoring in this cohort.

Left ventricular end diastolic pressure (LVEDP) is closely related to diastolic wall tension and is closely correlated with PCWP. In the Pexelizumab in Conjunction With Angioplasty in Acute Myocardial Infarction (APEX-AMI) study, an LVEDP greater than 22 mmHg was associated with higher rates of CS (4.6% vs 1.7%; P< 0.001) and death (4.1% vs 2.2%; P= 0.014) at 90 days.(21) We did not record LVEDP in our registry but PADP (but not PCWP) was associated with mortality at baseline and final datasets in both AMI-CS and HF-CS. Of note, PCWP data was only available in about one-third of patients while PADP was recorded in all patients, which may explain this discrepancy.

Baseline cardiac output was surprisingly lower in survivors than in non-survivors in the HF-CS cohort (4.0 ±1.4 vs 4.4± 1.6L/min, p=0.009), while on PAC removal it was higher in survivors (5.3±1.1 vs 4.7±1.6 L/min, p=0.001). This paradoxical finding could potentially be explained if patients with HF-CS with lower baseline CO were more likely to receive more aggressive or urgent treatment. Alternatively, patients with higher CO at PAC placement might have greater exposure to inotropes or MCS and experience treatment-related complications.

There are important limitations to our findings. First, we only include patients who received a PAC, which could introduce selection bias as patients with early shock may be managed without PAC, and patients with severe shock may die prior to PAC placement or treated with palliative measures. Second, criteria for use of PAC or treatment of CS, including the use of MCS, are not standardized. Thus, we cannot correlate hemodynamic trajectory with specific interventions. Third, in patients who had MCS before PAC insertion (31% HF-CS, 32.6% AMI-CS), baseline hemodynamics could underestimate the severity of CS at baseline. Similarly, the timing of final dataset could be data derived prior to proceeding with advanced options or death, which would significantly impact the data. Fourth, HF-CS was about twice as prevalent as AMI-CS (approximately two thirds to one third ratio). Hence, the analyses of the AMI-CS subset as compared to the HF-CS subset may be underpowered. Finally, the retrospective study design limits our ability to control for residual confounding.

## CONCLUSIONS

Data from a large, multicenter registry of patients with CS that undergo placement of a PAC indicate that clinical outcomes are not strongly associated with initial hemodynamics (especially in AMI-CS) which, in some cases can actually be misleading. This highlights the importance of monitoring and tracing the hemo-metabolic trajectory to tailor management. Further research is needed to define treatment pathways for improving hemodynamic and metabolic derangements in CS caused by HF and AMI.

## Data Availability

The data included in the manuscript is accurate and available

## Non-standard Abbreviations and Acronyms

AMI-CS: acute myocardial infarction cardiogenic shock
API: aortic pulsatility index
AUC: area under the curve
CI: cardiac index
CO: cardiac output
CPO: cardiac power output
CS: cardiogenic shock
CSWG: Cardiogenic Shock Working Group
CVP: central venous pressure
DBP: diastolic blood pressure
Delta-P: change in parameter
DPG: diastolic pulmonary gradient
ECMO: Extracorporeal membrane oxygenation
HF-CS: heart failure cardiogenic shock
HR: heart rate
HT: heart transplant
IABP: intra-aortic balloon pump
LVAD: left ventricular assist device
MAP: mean arterial pressure
MCS: mechanical circulatory support
PAC: pulmonary artery catheter
PADP: pulmonary artery diastolic pressure
PAPi: pulmonary artery pulsatility index
PCWP: pulmonary capillary wedge pressure
PVR: pulmonary vascular resistance
RAP: right atrial pressure
RVSWI: right ventricular stroke work index
SCAI: Society for Cardiovascular Angiography and Intervention
SBP: systolic blood pressure
SV: stroke volume
SVI: stroke volume index
SVR: systemic vascular resistance
TPG: transpulmonary gradient

## SUPPLEMENTAL MATERIALS (Intended for publication)

### Supplement Tables

**Supplement Table 1.** Formulas and Ranges of different Hemodynamic Parameters.

|  | Formulas | Normal Ranges |
| --- | --- | --- |
| Right Sided Parameters |  |  |
| RAP (mmHg) | - | 0-8 |
| PASP (mmHg) | - | 15-30 |
| mPAP (mmHg) | $2/3 \text{ PADP} + 1/3 \text{ PASP}$ | 8-20 |
| PAPi | $(\text{PASP}-\text{PADP})/\text{RAP}$ | > 2.5 (<1 indicative of RVF) |
| PVR (Woods Units) | $(\text{mPAP}-\text{PCWP})/\text{CO}$ | < 3 |
| PA Saturation (%) | - | 70-80 |
| PA Compliance (ml/mmHg) | $\text{SV}/(\text{PASP}-\text{PADP})$ | 3.8-12 (<2.5 indicative of RVF) |
| PA Pulse Pressure (mmHg) | $\text{PASP}-\text{PADP}$ | 7-19 |
| RVSWI (g/m/beat/m<sup>2</sup>) | $0.0136 * \text{SVI} * (\text{mPAP}-\text{RAP})$ | 5-10 (<5 indicative of worse outcome in HF) |
| RAP/PCWP ratio | $\text{RAP}/\text{PCWP}$ | <0.86 |
| RAP/PADP ratio | $\text{RAP}/\text{PADP}$ | <0.86 |
| TPG (mmHg) | $\text{mPAP}-\text{PCWP}$ | <12 |
| DPG (mmHg) | $\text{PADP}-\text{PCWP}$ | 0-5 |
| Left Sided Parameters |  |  |
| PADP (mmHg) | - | 4-12 |
| PCWP (mmHg) | - | 4-12 |
| CO (L/min) | - | 4-8 |
| CI (L/min/m<sup>2</sup>) | $\text{CO}/\text{BSA}$ | 2.5-4.2 |
| CPO (Watts) | $\text{MAP} * \text{CO}/451$ | >1 (<0.6 indicative of high mortality) |
| CPI W/m<sup>2</sup> | $\text{MAP} * \text{CI}/451$ | 0.5-0.7 |
| SVR (dynes.sec.cm<sup>-5</sup>) | $80 * (\text{MAP}-\text{RAP})/\text{CO}$ | 800-1200 |
| API | $(\text{SBP}-\text{DBP})/\text{PCWP}$ | 3-5 (<1.9 predictor of mortality in HF) |

Abbreviations like abbreviations for table 4.

**Supplement Table 2.** Baseline and Final Hemo-Metabolic Parameters Comparing AMI-CS and HF-CS.

|  | Baseline Dataset |  |  | Final Dataset |  |  |
| --- | --- | --- | --- | --- | --- | --- |
|  | AMI-CS<br>(N=604) | HF-CS<br>(n=1386) | p-values | AMI-CS<br>(N=604) | HF-CS<br>(n=1386) | p-values |
|  | Mean±SD | Mean±SD |  | Mean±SD | Mean±SD |  |
| Vital Signs |  |  |  |  |  |  |
| SBP (mmHg) | 109.7±22.1 | 107.9±19.1 | 0.15 | 106.6±23.3 | 106.5±23.1 | 0.94 |
| DBP (mmHg) | 66.3±16.7 | 67.9±13.9 | 0.07 | 59.4±15.2 | 64.1±13.7 | <0.001 |
| Heart rate (bpm) | 89.1±21.6 | 92.1±21.1 | 0.02 | 88.3±19.6 | 92.3±18.9 | <0.001 |
| Metabolic Parameters |  |  |  |  |  |  |
| Lactate (mmol/L) | 4.7±4.6 | 3.9±3.7 | 0.03 | 4.9±5.4 | 3.4±4.5 | 0.07 |
| ALT (IU/L) | 422.5±970.3 | 314.1±796.8 | 0.004 | 327.9±782.1 | 202.9±522.6 | <0.001 |
| AST (IU/L) | 435.8±1057.9 | 346.6±941.5 | <0.001 | 360.6±1198.0 | 192.1±671.8 | <0.001 |
| SCr (mg/dL) | 1.7±1.3 | 2.0±1.5 | <0.001 | 1.6±1.0 | 1.7±1.2 | 0.27 |
| Bicarbonate (mg/dL) | 21.7±5.4 | 23.5±6.1 | <0.001 | 23.2±5.0 | 25.2±5.5 | <0.001 |
| Hemodynamic Parameters |  |  |  |  |  |  |
| Left Sided Hemodynamics |  |  |  |  |  |  |
| PCWP (mmHg) | 21.5±9.8 | 23.2±9.4 | 0.20 | 19.7±9.1 | 18.3±8.1 | 0.22 |
| PADP (mmHg) | 23.1±8.7 | 26.4±9.5 | <0.001 | 19.6±7.5 | 20.9±8.9 | 0.03 |
| CO (L/min) | 4.3±1.7 | 4.0±1.4 | 0.13 | 4.9±1.5 | 5.2±1.6 | 0.02 |
| Cardiac index (L/min/m²) | 2.2±0.8 | 2.0±0.7 | 0.01 | 2.5±0.7 | 2.6±0.8 | 0.35 |
| CPO (Watts) | 0.8±0.3 | 0.7±0.3 | 0.14 | 0.8±0.3 | 0.9±0.3 | 0.04 |
| SVR (dynes.sec.cm<sup>-5</sup>) | 1471.6±637.6 | 1516.7±636.0 | 0.26 | 1150.3±473 | 1156.5±438.7 | 0.49 |
| API | 2.7±2.3 | 2.3±2.2 | 0.30 | 3.0±2.4 | 2.9±1.9 | 0.52 |
| Right Sided Hemodynamics |  |  |  |  |  |  |
| RAP (mmHg) | 13.4±6.3 | 14.4±7.3 | 0.04 | 10.9±5.5 | 10.7±6.4 | 0.23 |
| PASP (mmHg) | 42.9±14.7 | 50.7±16.3 | <0.001 | 40.2±13.2 | 43.8±15.5 | <0.001 |
| Mean PAP (mmHg) | 29.7±10.1 | 34.5±11.0 | <0.001 | 26.5±8.6 | 28.6±10.3 | 0.004 |
| PAPi | 2.0±1.9 | 2.5±2.9 | 0.003 | 2.7±2.8 | 3.4±3.6 | 0.002 |
| PVR (Woods Units) | 2.6±1.1 | 3.5±2.4 | 0.06 | 2.6±1.4 | 2.9±1.8 | 0.41 |
| PA Saturation (%) | 61.8±15.7 | 56.8±15.8 | 0.03 | 64±15.8 | 61.3±11.9 | 0.16 |
| PA Compliance (ml/mmHg) | 2.8±2.1 | 2.5±4.6 | <0.001 | 3.5±3.0 | 3.0±2.0 | 0.047 |
| RVSWI (g/m/beat/m2) | 6.0±3.5 | 6.9±15.5 | 0.25 | 6.9±3.4 | 7.2±4.0 | 0.78 |
| RAP/PCWP ratio | 0.6±0.2 | 0.6±0.4 | 0.10 | 0.6±0.3 | 0.6±0.3 | 0.96 |
| TPG (mmHg) | 10.0±8.5 | 11.0±8.3 | 0.58 | 9.0±7.8 | 10.2±6.9 | 0.10 |
| DPG (mmHg) | 1.9±8.3 | 2.3±7.3 | 0.64 | 0.5±6.7 | 2.2±6.3 | 0.05 |

AMI-CS indicates cardiogenic shock related to acute myocardial infarction; HF-CS, cardiogenic shock related to heart failure. The rest of the abbreviations are similar to abbreviations for table 4.

**Supplement Table 3.** Comparison of Baseline and Final Datasets Between Survivors and Non-survivors in HF-CS.

| Mean | Survivor (N=1070) |  |  | Non-survivor (N=311) |  |  |
| --- | --- | --- | --- | --- | --- | --- |
|  | Baseline | Final | p-value | Baseline | Final | p-value |
| **Metabolic Parameters** |  |  |  |  |  |  |
| Lactate (mmol/L) | 3.2 ± 2.8 | 1.8 ± 1.7 | <0.001 | 4.7 ± 4.6 | 5.4 ± 6.0 | 0.52 |
| BUN (mg/dL) | 38.6 ± 24 | 35.8 ± 22.2 | <0.001 | 46.1 ± 25.7 | 45.3 ± 27.4 | 0.70 |
| Serum Creatinine (mg/dL) | 1.9 ± 1.6 | 1.6 ± 1.2 | <0.001 | 2.2 ± 1.2 | 2.1 ± 1.2 | 0.28 |
| HCO<sub>3</sub> (mg/dL) | 24.1 ± 5.6 | 26.1 ± 4.6 | <0.001 | 21.9 ± 7.1 | 22.9 ± 6.8 | 0.07 |
| AST (IU/L) | 356 ± 1023 | 99 ± 340 | <0.001 | 315 ± 565 | 523 ± 1229 | 0.33 |
| **Vital Signs** |  |  |  |  |  |  |
| HR (bpm) | 91.4 ± 20.4 | 92.3 ± 17.5 | 0.19 | 94.8 ± 23.4 | 92.3 ± 23.5 | 0.18 |
| SBP (mmHg) | 108.7 ± 18.5 | 110.9 ± 20.7 | 0.01 | 105.3 ± 21.0 | 90.8 ± 24.3 | <0.001 |
| DBP (mmHg) | 69.1 ± 13.4 | 66.7 ± 12.4 | <0.001 | 63.9 ± 15.0 | 55.0 ± 14.4 | <0.001 |
| MAP (mmHg) | 82.3 ± 13.9 | 80.5 ± 12.9 | 0.003 | 77.5 ± 14.2 | 66.3 ± 16.1 | <0.001 |
| **Hemodynamic Parameters** |  |  |  |  |  |  |
| **Right Sided Hemodynamics** |  |  |  |  |  |  |
| RAP (mmHg) | 14.2 ± 7.3 | 9.7 ± 5.6 | <0.001 | 15.4 ± 7.5 | 14.3 ± 7.6 | 0.09 |
| PASP (mmHg) | 50.1 ± 15.7 | 43.1 ± 14.7 | <0.001 | 53.0 ± 18.0 | 46.2 ± 17.7 | <0.001 |
| mPAP (mmHg) | 34.1 ± 10.7 | 27.9 ± 10.0 | <0.001 | 36 ± 11.7 | 31.0 ± 11.1 | <0.001 |
| PAPi | 2.5 ± 3.0 | 3.6 ± 3.7 | <0.001 | 2.3 ± 2.6 | 2.4 ± 3.3 | 0.95 |
| PVR (Woods Units) | 3.4 ± 2.1 | 2.7 ± 1.4 | <0.001 | 4.1 ± 3.5 | 3.8 ± 3.2 | 0.26 |
| PA Saturation (%) | 56.6 ± 15.2 | 62.6 ± 11.1 | <0.001 | 58.1 ± 18.5 | 54.2 ± 13.8 | 0.56 |
| PA Compliance (ml/mmHg) | 2.3 ± 1.7 | 3.1 ± 2.0 | <0.001 | 3.4 ± 10.8 | 2.5 ± 2.0 | 0.14 |
| PA Pulse Pressure (mmHg) | 24.1 ± 10.8 | 23.1 ± 10.0 | 0.007 | 25.6 ± 12.4 | 22.8 ± 12.5 | 0.001 |
| RVSWI | 6.2 ± 3.4 | 7.3 ± 3.9 | <0.001 | 10.6 ± 37.5 | 6.7 ± 4.3 | 0.34 |
| RAP/PCWP ratio | 0.6 ± 0.4 | 0.6 ± 0.3 | 0.24 | 0.7 ± 0.4 | 0.7 ± 0.4 | 0.84 |
| RAP/PADP ratio | 0.6 ± 0.4 | 0.5 ± 0.3 | <0.001 | 0.6 ± 0.4 | 0.6 ± 0.3 | 0.66 |
| TPG (mmHg) | 10.5 ± 7.5 | 10.0 ± 6.5 | 0.35 | 14.5 ± 11.8 | 11.4 ± 9.1 | 0.07 |
| DPG (mmHg) | 1.8 ± 6.3 | 2.2 ± 5.8 | 0.37 | 5.3 ± 11.7 | 2.4 ± 8.8 | 0.45 |
| **Left Sided Hemodynamics** |  |  |  |  |  |  |
| PADP (mmHg) | 26.1 ± 9.4 | 20.2 ± 8.7 | <0.001 | 27.5 ± 9.8 | 23.5 ± 9.0 | <0.001 |
| PCWP (mmHg) | 23.5 ± 9.4 | 17.9 ± 7.7 | <0.001 | 21.4 ± 8.8 | 20.6 ± 10.3 | 0.28 |
| CO (L/min) | 4.0 ± 1.4 | 5.3 ± 1.6 | <0.001 | 4.4 ± 1.6 | 4.7 ± 1.6 | 0.06 |
| CI (L/min/m<sup>2</sup>) | 2.0 ± 0.7 | 2.7 ± 0.8 | <0.001 | 2.1 ± 0.7 | 2.3 ± 0.8 | 0.02 |
| CPO (Watts) | 0.7 ± 0.3 | 0.9 ± 0.3 | <0.001 | 0.8 ± 0.3 | 0.7 ± 0.3 | 0.42 |
| CPI (Watts/m<sup>2</sup>) | 0.4 ± 0.1 | 0.5 ± 0.2 | <0.001 | 0.4 ± 0.1 | 0.4 ± 0.1 | 0.69 |
| SVR (dynes.sec.cm<sup>-5</sup>) | 1549 ± 621 | 1175 ± 431 | <0.001 | 1346 ± 691 | 1058 ± 466 | <0.001 |
| API | 2.3 ± 2.3 | 2.9 ± 1.9 | <0.001 | 2.2 ± 1.5 | 2.4 ± 1.8 | 0.55 |

**Supplement Table 4.**
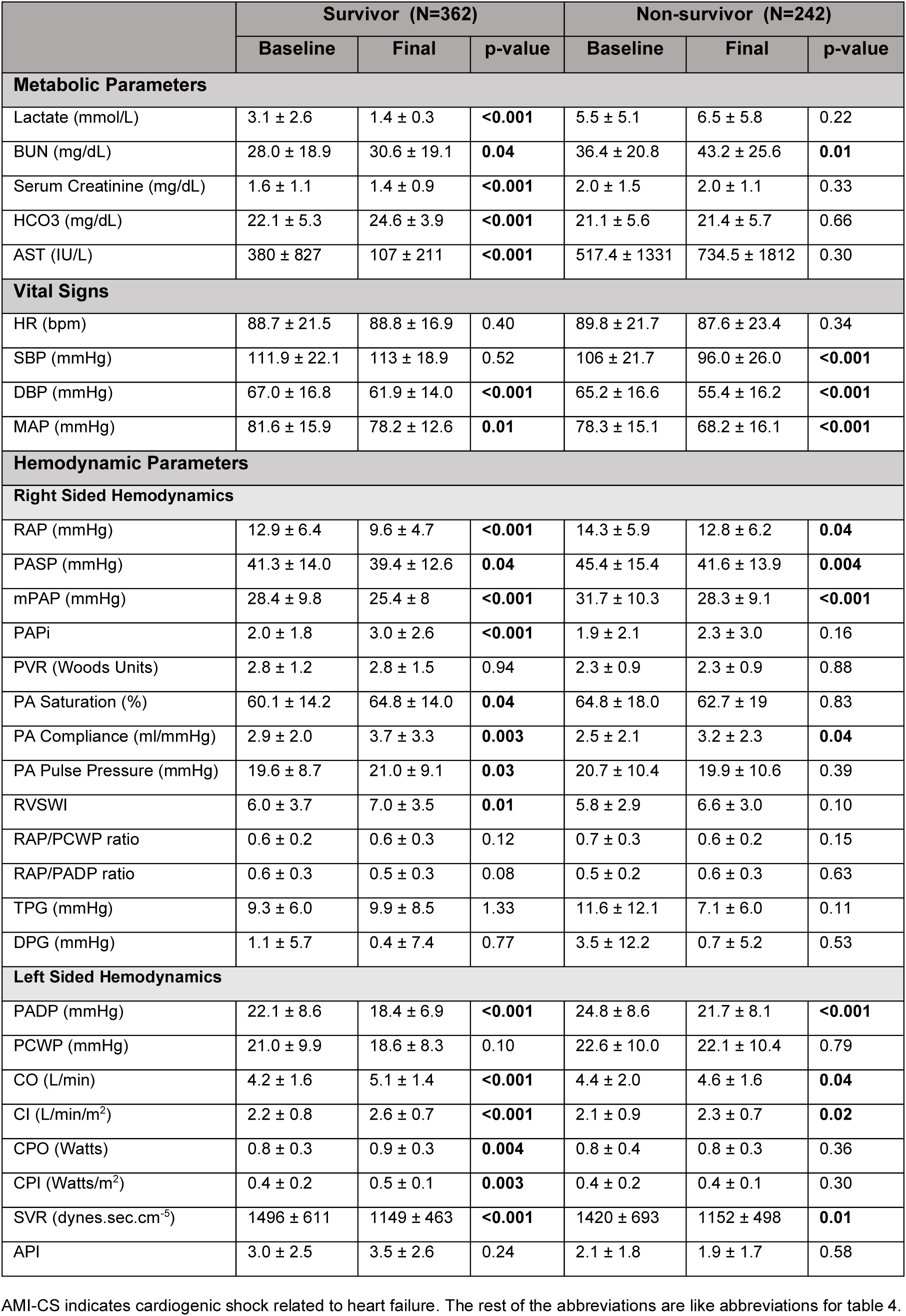
Comparison of Baseline and Final Datasets Between Survivors and Non-survivors in AMI-CS.

|  | Survivor (N=362) |  |  | Non-survivor (N=242) |  |  |
| --- | --- | --- | --- | --- | --- | --- |
|  | Baseline | Final | p-value | Baseline | Final | p-value |
| **Metabolic Parameters** |  |  |  |  |  |  |
| Lactate (mmol/L) | 3.1 ± 2.6 | 1.4 ± 0.3 | <0.001 | 5.5 ± 5.1 | 6.5 ± 5.8 | 0.22 |
| BUN (mg/dL) | 28.0 ± 18.9 | 30.6 ± 19.1 | 0.04 | 36.4 ± 20.8 | 43.2 ± 25.6 | 0.01 |
| Serum Creatinine (mg/dL) | 1.6 ± 1.1 | 1.4 ± 0.9 | <0.001 | 2.0 ± 1.5 | 2.0 ± 1.1 | 0.33 |
| HCO<sub>3</sub> (mg/dL) | 22.1 ± 5.3 | 24.6 ± 3.9 | <0.001 | 21.1 ± 5.6 | 21.4 ± 5.7 | 0.66 |
| AST (IU/L) | 380 ± 827 | 107 ± 211 | <0.001 | 517.4 ± 1331 | 734.5 ± 1812 | 0.30 |
| **Vital Signs** |  |  |  |  |  |  |
| HR (bpm) | 88.7 ± 21.5 | 88.8 ± 16.9 | 0.40 | 89.8 ± 21.7 | 87.6 ± 23.4 | 0.34 |
| SBP (mmHg) | 111.9 ± 22.1 | 113 ± 18.9 | 0.52 | 106 ± 21.7 | 96.0 ± 26.0 | <0.001 |
| DBP (mmHg) | 67.0 ± 16.8 | 61.9 ± 14.0 | <0.001 | 65.2 ± 16.6 | 55.4 ± 16.2 | <0.001 |
| MAP (mmHg) | 81.6 ± 15.9 | 78.2 ± 12.6 | 0.01 | 78.3 ± 15.1 | 68.2 ± 16.1 | <0.001 |
| **Hemodynamic Parameters** |  |  |  |  |  |  |
| **Right Sided Hemodynamics** |  |  |  |  |  |  |
| RAP (mmHg) | 12.9 ± 6.4 | 9.6 ± 4.7 | <0.001 | 14.3 ± 5.9 | 12.8 ± 6.2 | 0.04 |
| PASP (mmHg) | 41.3 ± 14.0 | 39.4 ± 12.6 | 0.04 | 45.4 ± 15.4 | 41.6 ± 13.9 | 0.004 |
| mPAP (mmHg) | 28.4 ± 9.8 | 25.4 ± 8 | <0.001 | 31.7 ± 10.3 | 28.3 ± 9.1 | <0.001 |
| PAPi | 2.0 ± 1.8 | 3.0 ± 2.6 | <0.001 | 1.9 ± 2.1 | 2.3 ± 3.0 | 0.16 |
| PVR (Woods Units) | 2.8 ± 1.2 | 2.8 ± 1.5 | 0.94 | 2.3 ± 0.9 | 2.3 ± 0.9 | 0.88 |
| PA Saturation (%) | 60.1 ± 14.2 | 64.8 ± 14.0 | 0.04 | 64.8 ± 18.0 | 62.7 ± 19 | 0.83 |
| PA Compliance (ml/mmHg) | 2.9 ± 2.0 | 3.7 ± 3.3 | 0.003 | 2.5 ± 2.1 | 3.2 ± 2.3 | 0.04 |
| PA Pulse Pressure (mmHg) | 19.6 ± 8.7 | 21.0 ± 9.1 | 0.03 | 20.7 ± 10.4 | 19.9 ± 10.6 | 0.39 |
| RVSWI | 6.0 ± 3.7 | 7.0 ± 3.5 | 0.01 | 5.8 ± 2.9 | 6.6 ± 3.0 | 0.10 |
| RAP/PCWP ratio | 0.6 ± 0.2 | 0.6 ± 0.3 | 0.12 | 0.7 ± 0.3 | 0.6 ± 0.2 | 0.15 |
| RAP/PADP ratio | 0.6 ± 0.3 | 0.5 ± 0.3 | 0.08 | 0.5 ± 0.2 | 0.6 ± 0.3 | 0.63 |
| TPG (mmHg) | 9.3 ± 6.0 | 9.9 ± 8.5 | 1.33 | 11.6 ± 12.1 | 7.1 ± 6.0 | 0.11 |
| DPG (mmHg) | 1.1 ± 5.7 | 0.4 ± 7.4 | 0.77 | 3.5 ± 12.2 | 0.7 ± 5.2 | 0.53 |
| **Left Sided Hemodynamics** |  |  |  |  |  |  |
| PADP (mmHg) | 22.1 ± 8.6 | 18.4 ± 6.9 | <0.001 | 24.8 ± 8.6 | 21.7 ± 8.1 | <0.001 |
| PCWP (mmHg) | 21.0 ± 9.9 | 18.6 ± 8.3 | 0.10 | 22.6 ± 10.0 | 22.1 ± 10.4 | 0.79 |
| CO (L/min) | 4.2 ± 1.6 | 5.1 ± 1.4 | <0.001 | 4.4 ± 2.0 | 4.6 ± 1.6 | 0.04 |
| CI (L/min/m<sup>2</sup>) | 2.2 ± 0.8 | 2.6 ± 0.7 | <0.001 | 2.1 ± 0.9 | 2.3 ± 0.7 | 0.02 |
| CPO (Watts) | 0.8 ± 0.3 | 0.9 ± 0.3 | 0.004 | 0.8 ± 0.4 | 0.8 ± 0.3 | 0.36 |
| CPI (Watts/m<sup>2</sup>) | 0.4 ± 0.2 | 0.5 ± 0.1 | 0.003 | 0.4 ± 0.2 | 0.4 ± 0.1 | 0.30 |
| SVR (dynes.sec.cm<sup>-5</sup>) | 1496 ± 611 | 1149 ± 463 | <0.001 | 1420 ± 693 | 1152 ± 498 | 0.01 |
| API | 3.0 ± 2.5 | 3.5 ± 2.6 | 0.24 | 2.1 ± 1.8 | 1.9 ± 1.7 | 0.58 |

AMI-CS indicates cardiogenic shock related to heart failure. The rest of the abbreviations are like abbreviations for table 4.

**Supplement Table 5.** Univariable Logistic Regression of Hemo-Metabolic and Variables for Mortality Association in the Total Cohort.

| Variable | Baseline Dataset |  | Final Dataset |  | Delta |  |
| --- | --- | --- | --- | --- | --- | --- |
|  | Odds ratio<br>(Lower-Upper CL) | p-value | Odds ratio<br>(Lower-Upper CL) | p-value | Odds ratio<br>(Lower-Upper CL) | p-value |
| **SBP** | 0.99 (0.98-1) | **0.02** | **0.96 (0.95-0.96)** | **<0.0001** | **0.97 (0.97-0.98)** | **<0.0001** |
| **DBP** | 0.97 (0.96-0.98) | **<0.0001** | **0.93 (0.92-0.94)** | **<0.0001** | **0.98 (0.97-0.99)** | **<0.0001** |
| **HR** | 1.01 (1-1.01) | **0.03** | **1 (0.99-1.01)** | 0.98 | 0.99 (0.99-1) | **0.04** |
| **PCWP** | 0.98 (0.94-1.01) | 0.19 | 1.04 (1-1.08) | **0.04** | **1.05 (1.01-1.08)** | **0.01** |
| **PADP** | 1.01 (1-1.03) | **0.05** | **1.04 (1.02-1.06)** | **<0.0001** | **1.02 (1-1.03)** | **0.02** |
| **CO** | 1.21 (1.07-1.38) | **<0.01** | **0.80 (0.70-0.91)** | **<0.01** | **0.74 (0.66-0.83)** | **<0.0001** |
| **CI** | 1.37 (1.04-1.8) | **0.02** | **0.58 (0.44-0.77)** | **<0.01** | **0.56 (0.44-0.70)** | **<0.0001** |
| **CPO** | 2.01 (0.98-4.11) | **0.06** | **0.12 (0.06-0.27)** | **<0.0001** | **0.13 (0.07-0.25)** | **<0.0001** |
| **RAP** | 1.02 (1-1.04) | **0.04** | **1.11 (1.09-1.14)** | **<0.0001** | **1.06 (1.03-1.08)** | **<0.0001** |
| **PASP** | 1.01 (1-1.02) | **0.02** | **1.01 (1-1.02)** | **0.01** | **1 (0.99-1.01)** | 0.86 |
| **MPAP** | 1.02 (1-1.03) | **0.02** | **1.03 (1.01-1.04)** | **<0.0001** | **1.01 (1-1.02)** | 0.16 |
| **PAPI** | 0.97 (0.91-1.04) | 0.36 | 0.85 (0.78-0.92) | **<0.01** | **0.94 (0.9-0.98)** | **0.01** |
| **PA SAT** | 1.01 (0.98-1.03) | 0.62 | 0.94 (0.91-0.97) | **<0.01** | **0.97 (0.95-0.99)** | **<0.01** |
| **RVSWI** | 1.06 (1-1.12) | **0.06** | **0.96 (0.91-1.02)** | 0.19 | 0.93 (0.88-0.98) | **0.01** |
| **TPG** | 1.05 (1.01-1.09) | **0.01** | **1.03 (0.98-1.07)** | 0.26 | 0.96 (0.92-1) | **0.03** |

Abbreviations like abbreviations for table 4.

